# Partnering Early to Provide for Infants At Risk of Cerebral palsy (PĒPI ARC): Protocol for a feasibility study of a regional hub for early detection of cerebral palsy in Aotearoa New Zealand

**DOI:** 10.1101/2023.10.31.23297869

**Authors:** Angelica Allermo Fletcher, Gaela Kilgour, Meghan Sandle, Sally Kidd, Alison Sheppard, Stephanie Swallow, Ngaire Susan Stott, Malcolm Battin, Wyllis Korent, Sian A Williams

**Affiliations:** Neonatal Intensive Care Unit, Wellington Regional Hospital, Wellington, New Zealand; Wellington Regional Hospital; Child Development Service, Wellington Regional Hospital, Wellington, New Zealand; Newborn Service, Auckland City Hospital; School of Allied Health, Curtin University, Perth, Australia. University of Auckland, Liggins Institute, Auckland, New Zealand

## Abstract

**Introduction:** Cerebral palsy (CP) can now be diagnosed as early as three months of age in infants with identified CP risk factors, but many barriers prevent equitable access to early detection pathways. The “Partnering Early to Provide for Infants At Risk of Cerebral Palsy” feasibility study (PĒPI ARC) seeks to trial a new approach to decrease inequitable health service in Aotearoa New Zealand for high-risk infants and their families. PĒPI ARC will incorporate face-to-face clinics, an in-person and virtual Hub, and the use of telehealth to enable flexible access to CP assessments, and support for health professionals in early CP detection.

**Methods and Analysis:** A non-randomised feasibility study will be conducted from the tertiary Neonatal Intensive Care Unit in Wellington and includes seven regional referral centres, servicing nearly 30% of the NZ total population. Families of infants with high risk of neurodevelopmental impairment will be invited to participate, as well as health professionals interacting with the Hub. Mixed methods will be used to evaluate the i) equitable implementation of an early detection pathway, ii) acceptability, iii) demand among families and health professionals, iv) efficacy in relation to reducing the age of receipt of CP diagnosis, and v) the experiences around communication and information sharing.

**Ethics and Dissemination:** The New Zealand Health and Disability Ethics Committees approved this study (HDEC:2022 FULL 13434). Findings will be disseminated in peer-reviewed journals, conference presentations and via professional networks.

**Registration:** Australian New Zealand Clinical Trials Registry: ACTRN12623000600640

**STRENGTHS AND LIMITATIONS OF THE STUDY:**

- The New Zealand Best Practice Recommendations for early detection of CP are based on international guidelines and have been peer reviewed for the Aotearoa New Zealand context.
- Local and regional health professionals have collaborated to inform the PĒPI ARC protocol with the aim to improve access to early CP assessments and early detection rates of CP.
- Reduction in health inequities for Māori and Pasifika have been targeted through informed partnerships.
- Resource development and planning of PĒPI ARC Hub has been co-designed with families and recognises the ecological context of Aotearoa New Zealand.
- A limitation is that only high-risk infants with “newborn detectable risks” will be included in the study.

## INTRODUCTION

In Aotearoa New Zealand (NZ), it is estimated that every three days a baby is born who will later receive a diagnosis of Hōkai Nukurangi, the Te Reo Māori term for cerebral palsy (CP)[1]. Of these babies, approximately 60% will be diagnosed after 12 months of age, with a reported median age of CP diagnosis at 17 months of age in NZ [1]. Many families of children with CP living in NZ perceive diagnosis delays, with nearly 20% of families reporting delays of more than 12 months[2].

Historically, CP has been diagnosed using a combination of clinical and neurological signs, which often do not appear until the child is older. In 2017, an international evidenced-based guideline was published for accurate and early diagnosis of CP using a combination of different assessment tools[3]. By combining the Prechtl Quality Assessment of General Movements (GMA), Hammersmith Infant Neurological Examination (HINE) and brain Magnetic Resonance Imaging (MRI), infants with detectable newborn risk factors for CP can now be diagnosed as early as 3 months of age (corrected for prematurity) with a sensitivity of 97 %[4]. The successful implementation of these recommendations have seen significant reductions in the age of identifying ‘high risk of CP’ (e.g. in an Australia clinic with an average age of 4.4 months[5]) and the average age of CP diagnosis (e.g., in a USA clinic from 18 months prior to implementation down to 12 months[6], in a network of USA clinics 19.5 months to 9.5 months[7], and in an Australian clinic to 8.5 months[5]).

Based on international recommendations[3, 8], the New Zealand Cerebral Palsy Clinical Network (CPCN) in 2020 developed national *Best Practice Recommendations for Early Detection of CP, Intervention and Monitoring* (BPR) to support the implementation of early clinical CP detection pathways across NZ. The development of the BPR provides clear pathways[9] and guidance for consistent assessment approaches[10], but it does not remove other barriers to implementing early detection in practice. These barriers, as reported by health professionals, include lack of resources and multi-disciplinary teams[10], poor coordination between hospital, community and regional follow-up, and inequitable access to services[9–11]. Families report dissatisfaction and distrust with the diagnostic process, delays in diagnosis, and issues with communication and access to practical and emotional support[2]. In response to these concerns, co-design workshops for Māori and non-Māori families were conducted and revealed the need for improved service provision and navigation within a complex health system, with a ‘one-stop shop’ solution proposed as a more equitable and culturally safe system[12]. The PĒPI ARC Hub seeks to incorporate these learnings within the implementation of the BPR.

## P**Ē**PI ARC

The PĒPI ARC Hub applies a modified multidisciplinary team approach to health service delivery and care for infants with detectable risk factors for CP admitted to a tertiary NICU. The Hub aims to champion a Māori- centred relationship model of care with Whanaungatanga (connectedness) and Whakawhanaungatanga (building relationships) at its core[13], incorporating the key interrelated dimensions of Māori holistic health care including Wairua (spiritual), whānau (extended family network), hinengaro (the mind, emotion) and tinana (physical). Whānau represents the collective way Māori operate. People do not exist in isolation but include a person’s immediate and extended family. PĒPI ARC embraces the view that at the centre of care is the patient (i.e. infant) and their extended family and seeks to: 1) build relationships and connections through an infant’s journey in NICU, their transition to home and follow-up support in their community[14], and 2) coordinate care and minimise the burden for families to attend multiple appointments with different health professionals.

The Hub will provide three functions:

1. A face-to-face clinic for infants and their family living in Wellington.
2. Support to families and infants who reside in other regions to access GMA and HINE assessments, by working in close partnership with family, their local doctors, and therapists.
3. Partner with, and provide support for, local and regional health professionals to use early detection strategies and communication with families.

### Study aims and objectives

This study will evaluate the feasibility of equitable implementation of a pathway for the early detection of CP for high-risk infants (criteria outlined in Table 1) admitted to a tertiary NICU in NZ through a multidisciplinary regional Hub - The PĒPI ARC Hub. Based on the work of Bowen et al[15](i) **Implementation**, (ii) **Acceptability**[16] (iii) **Demand** and (iv) **Limited-Efficacy** of PĒPI ARC will be studied.

**Table 1.**
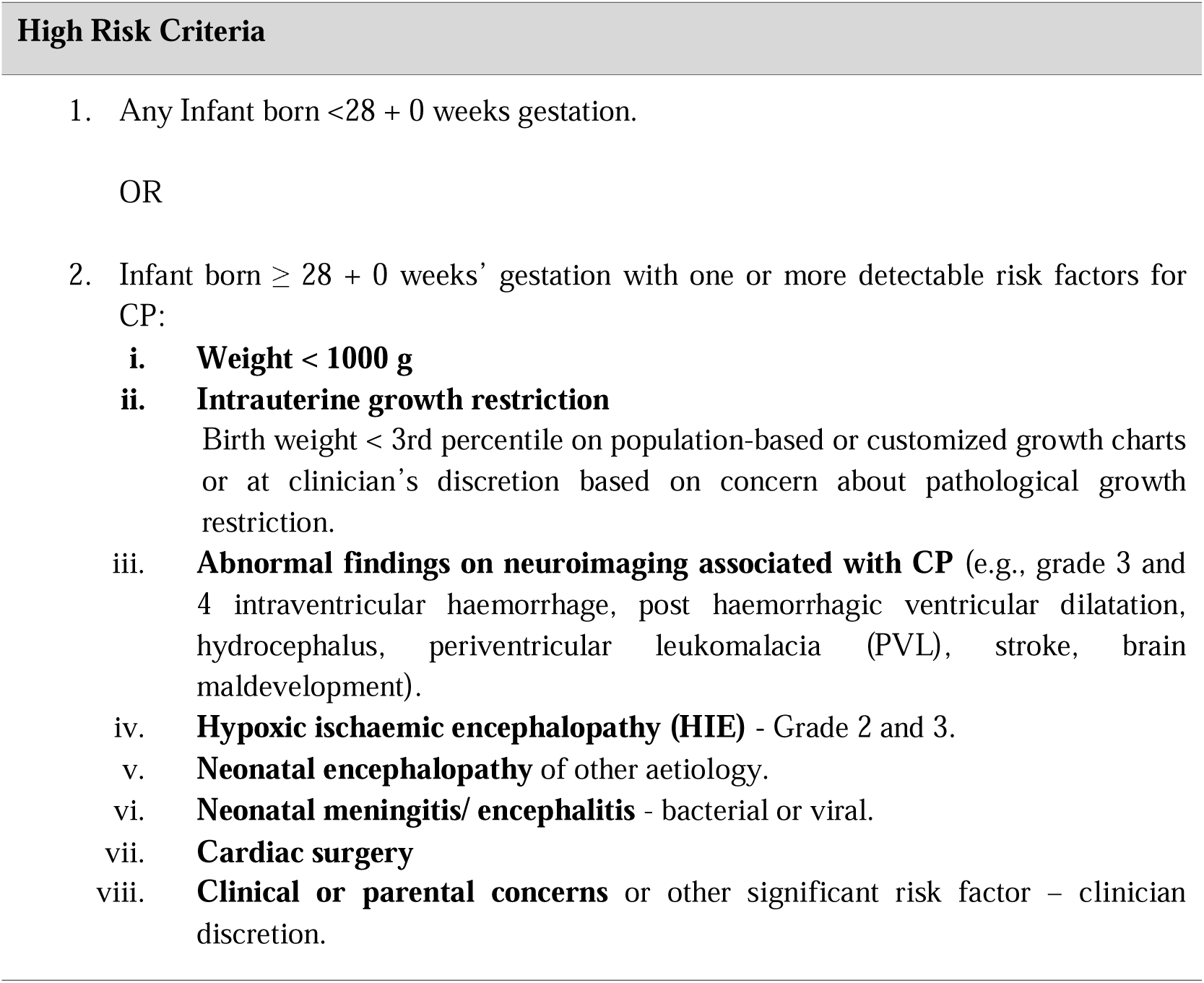
“High Risk” criteria for infants within the Newborn detectable risks pathway, as outlined in the NZ BPR for Early detection of cerebral palsy, intervention and monitoring.

### Primary Objective

1. **Implementation:** To determine to what extent the BPR for early diagnosis of CP can be equitably implemented for high-risk infants admitted to a level 3 NICU (independent of place of residence and ethnicity), and if the fidelity of the BPR can be maintained.

### Secondary Objectives

**2. Acceptability:** To evaluate the acceptability of a PĒPI ARC Hub model of care by families and health professionals.
**3. Demand:** To determine the demand for the PĒPI ARC Hub among both local and regional families and health professionals.
**4. Limited-Efficacy:**

i. To evaluate if the PĒPI ARC Hub model reduces the age of diagnosis of CP.
ii. To explore the experiences around communication and information sharing between families and health professionals.

## METHODS AND ANALYSIS

This study protocol was prepared according to the Standard Protocol Items: Recommendations for

Interventional Trials (SPIRIT) checklist[17], with adaptations based on recommendations outlined by Thebane and Lancaster[18].

### Study design and setting

The PĒPI ARC study is a prospective, non-randomised, feasibility study using a mixed methods approach to evaluate the previously outlined domains of feasibility. The study will run from March 2023-September 2024, with the primary time-point being at the completion of the PĒPI ARC Hub trial period. The study setting will be a regional Hub based in Wellington for follow-up of eligible infants who have received care at Wellington NICU. The PĒPI ARC Hub will be delivered using two components: a community based, multidisciplinary face-to-face diagnostic clinic in Wellington, and a virtual Hub to support regional CP-specific neurodevelopmental follow up, education and information sharing and foster relationships between the Wellington team, local community, and regional Child Development Services (CDS) and paediatricians.

Wellington NICU is a tertiary unit providing perinatal and surgical care for infants from Wellington and seven regional referral centres from half of the North Island and top of the South Island of NZ. The unit cares for a third of all infants admitted to a tertiary NICU in NZ. It is estimated that between 100-120 infants per year will fit the criteria for the CP BPR pathway, of which approximately 30% live in the Wellington area[19] (i.e. to be invited to attend the in-person clinic), and the remainder will receive follow-up in their local regions (supported by PĒPI ARC team through telehealth).

For the Wellington NICU catchment area, data from the New Zealand CP Register (2009-2020) showed that 17% of infants received a diagnosis of CP before 6 months of age and 34% before 12 months[20].

### Patient and Public Involvement

The development and refinement of the study question, study protocol, Hub design and resources were guided by three working groups: Whānau Hui, Advisory Group (provided local and cultural advice), and Steering Group. A specific focus was placed on the voice of Māori and Pasifika whānau, who are reported to have higher rates of preterm birth[21] and poorer preterm outcomes[22]. In the planning stages of PĒPI ARC, whānau hui (meetings/gathering with family members of infants who had previously been through the NICU) involved focused discussions about their Wellington NICU experience, reflections on their journey, use of assessment tools and possible improvements. Families informed the study design in the following areas:

- Early diagnosis conversations and information sharing- consistent, timely, honest, appropriate.
- Service Delivery: The experience of early diagnostic assessments in NICU and within the Community Service.
- Hearing the Parent’s Voice.
- Hub design.
- Cultural needs and safety.

The PĒPI ARC advisory group consists of families and health professionals including a Māori researcher and advisor to support our commitment to Te Tiriti O Waitangi and equitable health outcomes for Māori, and a Pasifika social worker. The PĒPI ARC Steering Group, formed by experts in early detection of CP from NZ and Australia, guided the planning and provide ongoing oversight of the study.

### Participants and recruitment

Participants for this study will be formed by two groupings, the infants and their parent/caregivers, and the health professionals involved/interacting with the PĒPI ARC Hub. There are no limits placed on sample size and a formal sample size calculation is not required. There will be no restrictions on other care or interventions partaken in by participants.

#### Infants and parents/ caregivers

Infants must meet the following eligibility criteria:

1. admitted to the NICU at Wellington Regional Hospital within the study period,
2. have a discharge address located within the Wellington region or within one of Wellington NICU’s referral regions,
3. and meet the BPR criteria for high risk of CP (Table 1).

Infants will be excluded if they are born with a life limiting condition and not expected to survive past the first year of life, or if the family is expected to move out of the catchment area before the infant is 3 months CGA.

Infants and their parent/caregivers will be invited to participate in the study by a member of the PĒPI ARC research team (not directly involved in providing health care for the infant in the NICU). Recruitment can occur at any point during the NICU admission prior to discharge. Support through social workers and Whānau Care Services is available to assist with recruitment decision making, and parents are encouraged to discuss and seek support from their extended family. Families will be advised that they can choose Usual care option (see below).

#### Health professionals

All health professionals who engage with the PĒPI ARC Hub will be invited to complete a questionnaire about their experience and acceptability of the PĒPI ARC model of care. There are no exclusion criteria. Health professionals will be invited to complete a semi-structured interview about their experience and acceptability of the PĒPI ARC Hub.

Members of the PĒPI ARC team will complete a team specific questionnaire focused on the acceptability of PĒPI ARC.

### Intervention - While in NICU

#### All infants enrolled in PĒPI ARC

All families will receive:

- *Welcome folder* with information about the study and the PĒPI ARC team, and documents to help them navigate their NICU journey and transfer to a regional centre or discharge home with community follow-up. They will also be provided with education and instructions on filming and transferring of GMA videos to the PĒPI ARC team after discharge.
- *PĒPI Passport:* aimed to improve health literacy, support a relationship model of care in the NICU, allow families to share neurodevelopmental information about their infant with different healthcare providers after discharge from NICU and to help smooth the transition from NICU to regional centres.

The passport was developed in partnership with health professionals and families and includes both English and Te Reo Māori translations. Families can document their infant’s developmental strengths and challenges, completed assessments and results, and plan for medical and developmental follow-up.

#### Infants and families living in the regional referral areas

A PĒPI ARC research nurse will support out-of-region families throughout their NICU journey, upon discharge/transfer and follow up with regional services to provide continuity of care as part of the ‘one stop shop’. Infants from Wellington will be supported by the Discharge facilitator team as part of NICU usual care.

### Intervention - After discharge from NICU

#### Infants and families living within the Wellington region Face-to-face clinic

A face-to-face multidisciplinary team (MDT) clinic appointment (lasting 90 minutes) will take place in a community setting when the infant is between 12-16 weeks CGA. This Hub appointment will replace their usual care appointment with their hospital neonatologist at the same age. The MDT will consist of a neonatologist or developmental paediatrician, a GMA and HINE trained therapist, a social worker, a clinic nurse, and may also include assessments by a Speech and Language Therapist (SLT)/Dietician if clinically indicated.

The infant will have a growth, development, medical and social work assessment, and The Feeding Matters Infant and Child Feeding Questionnaire (ICFQ)[23] evaluated. Any previously collected GMA and neuroimaging will be reviewed, and a HINE and GMA performed (Table 2). The family will also have time with the social worker in private. Assessment findings, resources and support will be shared with the family as part of the appointment (as outlined in Table 3), and with the infant’s local health care providers via an electronic assessment report. Both the family and health providers will also receive a detailed clinic letter within two weeks.

**Table 2.** PĒPI ARC clinic assessment schedule (12-14 weeks CGA)

| Assessment | Schedule | Trigger for second clinic visit/ referrals |
| --- | --- | --- |
| GMA | General Movement Assessment of presence or absence of normal fidgety movements.<br>Review of video captured by parents/caregivers prior to clinic, or video taken at clinic visit<sup>#</sup> | Repeated 2 weeks later if abnormal/absent |
| **HINE** | • 3 months Hammersmith Infant Neurological Examination | Repeated at 4 months CGA (16-18 weeks) if assessments are inconclusive at the initial appointment. |
| **Neuroimaging**<br>**HUS/MRI** | • Review of prior neuroimaging and need for further imaging assessed. | Referral for MRI (if not already obtained) if:<br>• abnormal/absent GMA, and/or<br>• HINE score (<57), and<br>• Assessed as high-risk of CP |
| **ICFQ<sup>[23]</sup>** | • Completed by the family before or at the clinic appointment | Referral to SLT and/ or dietician will be made (if not already involved) if two or more questions indicate red flags for feeding difficulties<sup>[23]</sup> |

<sup>#</sup>scored by the GMA team or at least two senior assessors (inter-rater reliability of the assessors will be established beyond 80% prior to PĒPI ARC clinic appointments beginning.) If consensus on scoring is not reached, the GMA will be reviewed independently by a third assessor, including referral to expert assessors if needed until consensus is reached.

**Table 3.**
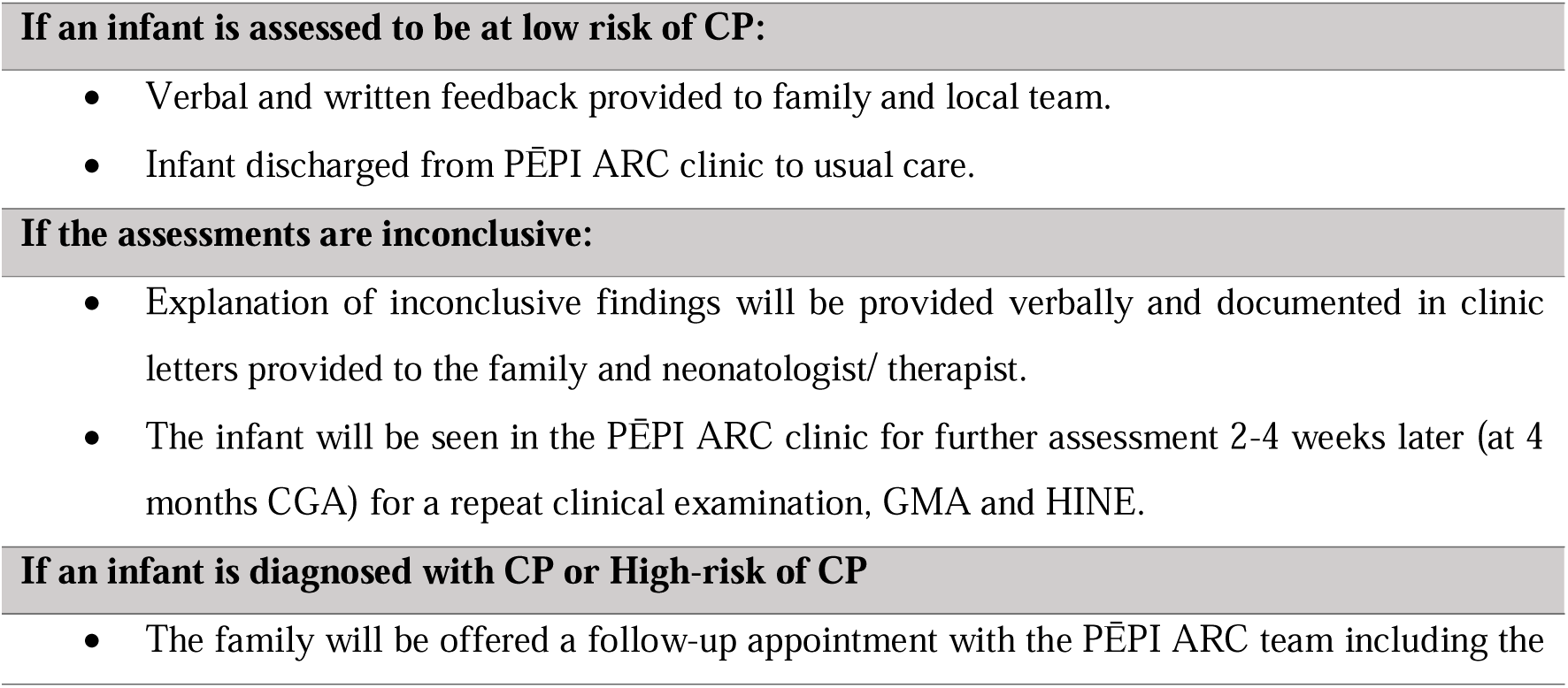
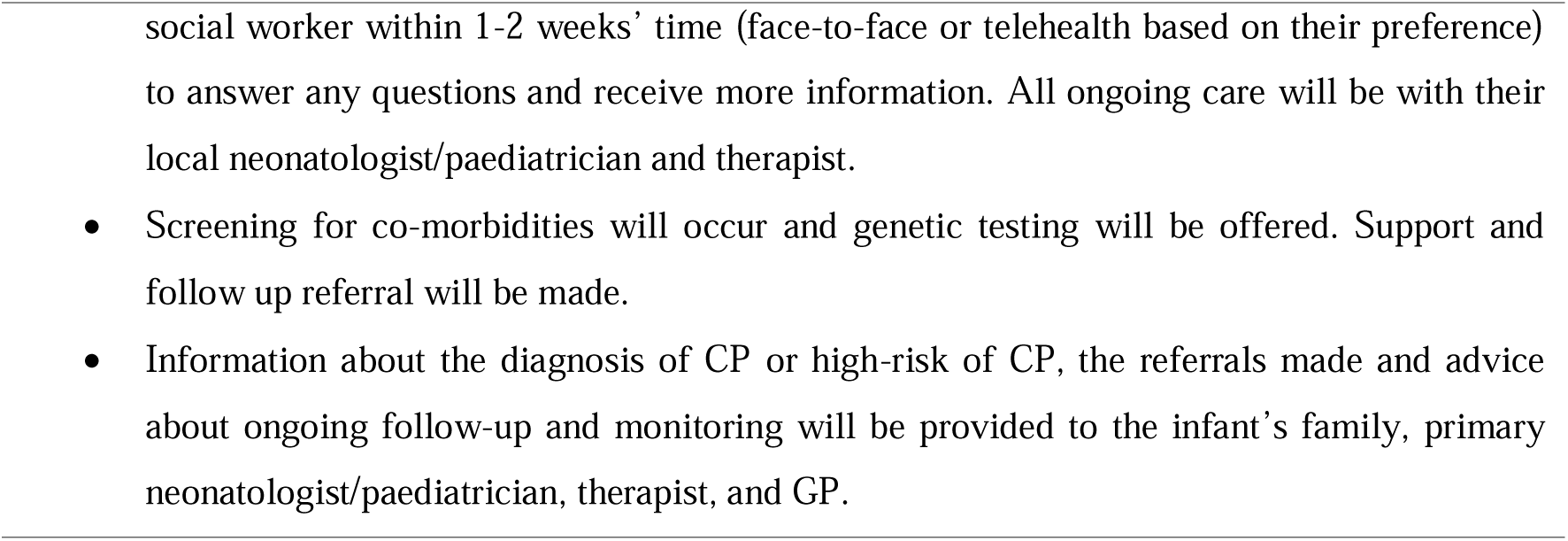
Process plan for communicating outcomes to families / health professionals.

#### Telehealth

Local families unable to attend the clinic in-person will be offered their MDT assessment using telehealth with assistance from the therapist or clinic nurse. They are also able to contact the Hub/ PĒPI ARC team via email and phone at any time during the study time to request support or answer questions.

#### Infants and families living outside of the Wellington region Virtual Hub

The virtual PĒPI ARC Hub will offer flexible support for families, paediatricians, and therapists to aid timely completion of early detection assessments (Table 2). The Hub will be able to assist with interpretation of specific assessments, triangulation of the results and help with CP diagnosis, and communicating assessment outcomes to families. If an infant receives a high risk of CP/CP diagnosis, additional information relating to early intervention, monitoring of comorbidities and complications of CP, parental support, and parent information about CP will be offered.

### Usual Care for infants at high risk of neurodevelopmental impairment admitted to Wellington NICU

#### All high-risk infants

- Routinely referred to the unit’s neonatal therapist, and on an as needed basis to other allied health.
- Whilst staff try to implement BPR, early GMA assessments are not always completed due to infant medical status and availability.
- MRI is routinely done only for infants with HIE grade 2 and 3.

#### Infants from Wellington region

- Followed by the Discharge facilitator team throughout their NICU admission and for a period after discharge. This team provides emotional and practical support, prepares families for discharge, organises family meetings, ensures follow-up appointments are made and assists families once home with feeding support including nasogastric tube feeding, and assessment of infants between clinic visits.
- Referred to a Visting Neonatal Therapist on discharge for neurodevelopmental surveillance, who may assist with GMA and HINE. The onset of this service and the length of follow-up time may vary.
- Followed-up by their hospital neonatologist, variable number of appointments and length of follow- up.
- Follow-up with SLT, dietician and other health professionals only if referred prior to discharge.
- No social work follow-up.
- No MDT clinics.

#### Infants from outside the Wellington region

- There is no service similar to the discharge facilitators team for infants whose families reside outside of Wellington during their NICU stay.
- Referred for paediatric and CDS for follow up and neurodevelopmental surveillance on discharge from NICU. Variation between regions for the onset of services, length of follow-up time, number of appointments and assessments[24].
- Three regional centres have clinicians that trained in GMAs, and a further three regional centres send GMA videos to the Wellington GMA team for scoring.
- An electronic report with individual GMA scores and an impression of outcome based on the trajectory at 3-4 months CGA is forwarded by the Wellinton GMA team to local therapist following evaluation of scores.
- No MDT clinics.

It is important to acknowledge that over the course of the study timeline, changes in ‘usual care’ may occur.

### Outcome measures and data collection

Feasibility will be assessed by using the outcomes, measures and data capture summarised in Table 4, from March 2023 to September 2024. Data will be directly entered into REDCap (hosted by Wellington Hospital and MRINZ infrastructures) which will only be accessed by members of the research team involved in data entry and analyse. Due to the scope and minimal risks associated with the study, there is no formal data monitoring committee. Interim data monitoring will be conducted at a 6-month review of survey feedback, and auditing of trial conduct will be completed by the primary investigator, and study and clinical coordinators on a fortnightly basis. Quantitative data collection tools, surveys for families, health professionals and the PĒPI ARC MDT team, participant information, consent form and Data management plan are available on Open Science Framework (OSF).

**Table 4.**
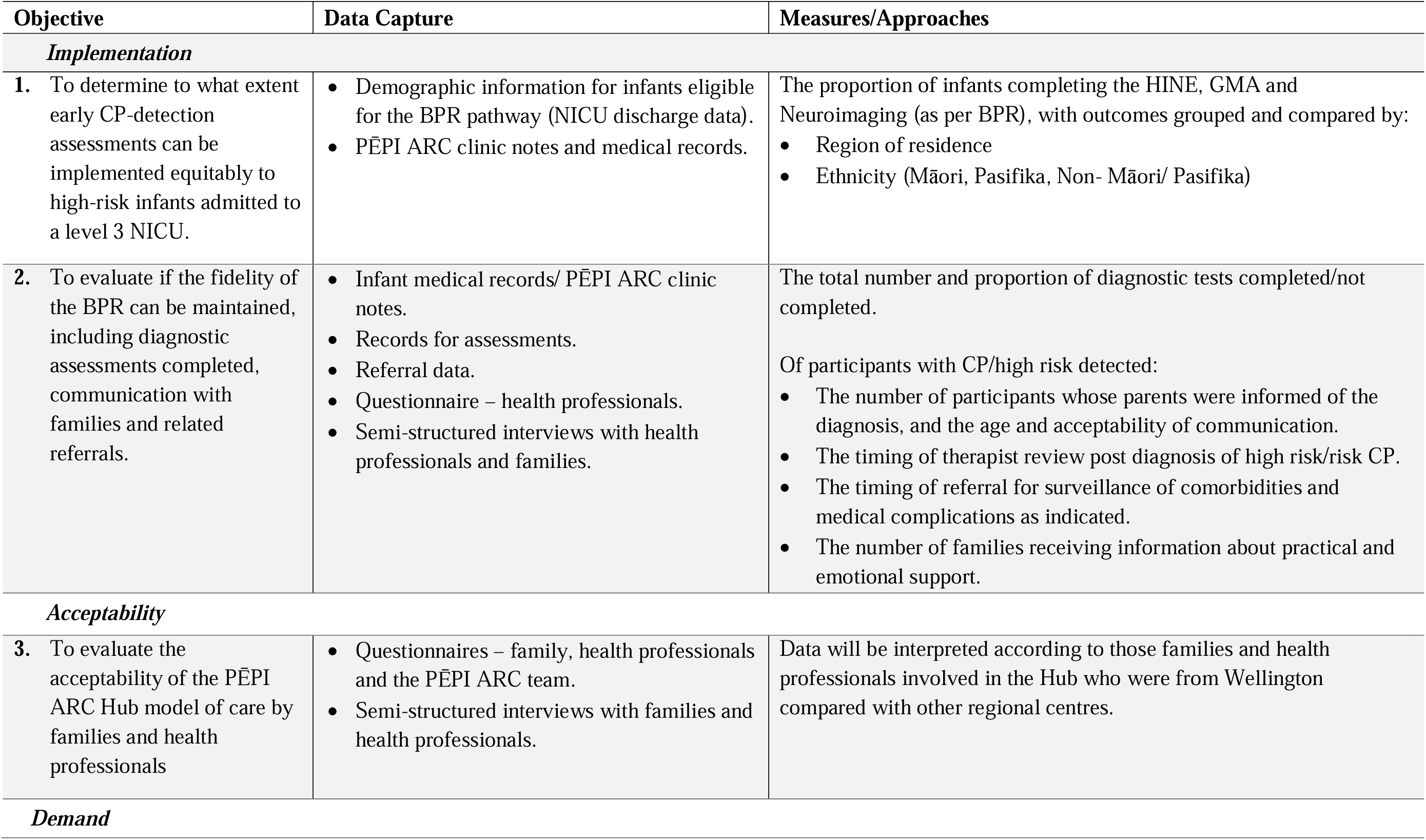
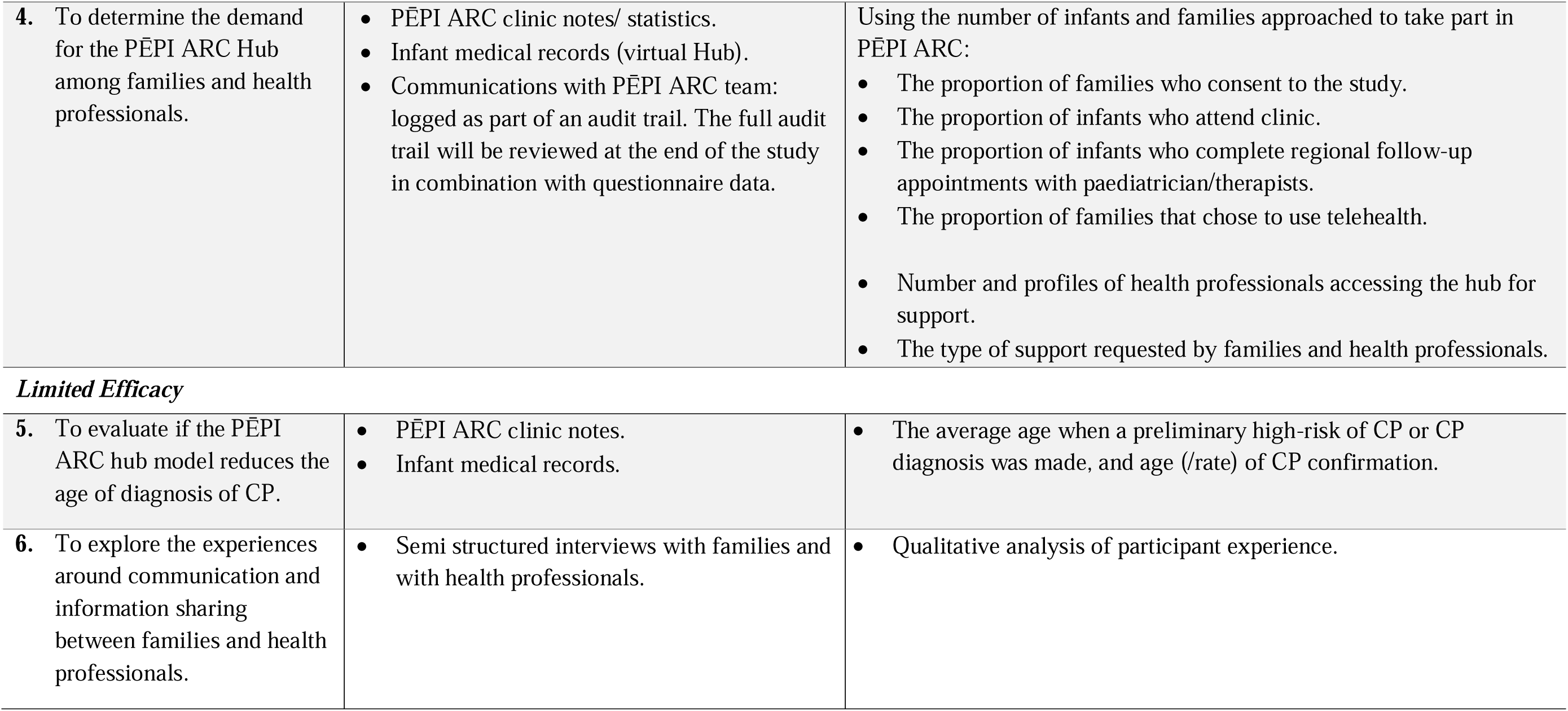
Summary of Outcomes, Data Capture and Measures/Approaches for PĒPI ARC.

| Objective | Data Capture | Measures/Approaches |
| --- | --- | --- |
| **Implementation** |  |  |
| 1. To determine to what extent early CP-detection assessments can be implemented equitably to high-risk infants admitted to a level 3 NICU. | Demographic information for infants eligible for the BPR pathway (NICU discharge data).<br>PĒPI ARC clinic notes and medical records. | The proportion of infants completing the HINE, GMA and Neuroimaging (as per BPR), with outcomes grouped and compared by:<br>Region of residence<br>Ethnicity (Māori, Pasifika, Non- Māori/ Pasifika) |
| 2. To evaluate if the fidelity of the BPR can be maintained, including diagnostic assessments completed, communication with families and related referrals. | Infant medical records/ PĒPI ARC clinic notes.<br>Records for assessments.<br>Referral data.<br>Questionnaire – health professionals.<br>Semi-structured interviews with health professionals and families. | The total number and proportion of diagnostic tests completed/not completed.<br>Of participants with CP/high risk detected:<br>The number of participants whose parents were informed of the diagnosis, and the age and acceptability of communication.<br>The timing of therapist review post diagnosis of high risk/risk CP.<br>The timing of referral for surveillance of comorbidities and medical complications as indicated.<br>The number of families receiving information about practical and emotional support. |
| **Acceptability** |  |  |
| 3. To evaluate the acceptability of the PĒPI ARC Hub model of care by families and health professionals | Questionnaires – family, health professionals and the PĒPI ARC team.<br>Semi-structured interviews with families and health professionals. | Data will be interpreted according to those families and health professionals involved in the Hub who were from Wellington compared with other regional centres. |
| **Demand** |  |  |
| 4. To determine the demand for the PĒPI ARC Hub among families and health professionals. | • PĒPI ARC clinic notes/ statistics.<br>• Infant medical records (virtual Hub).<br>• Communications with PĒPI ARC team: logged as part of an audit trail. The full audit trail will be reviewed at the end of the study in combination with questionnaire data. | Using the number of infants and families approached to take part in PĒPI ARC:<br>• The proportion of families who consent to the study.<br>• The proportion of infants who attend clinic.<br>• The proportion of infants who complete regional follow-up appointments with paediatrician/therapists.<br>• The proportion of families that chose to use telehealth.<br>• Number and profiles of health professionals accessing the hub for support.<br>• The type of support requested by families and health professionals. |
| *Limited Efficacy* |  |  |
| 5. To evaluate if the PĒPI ARC hub model reduces the age of diagnosis of CP. | • PĒPI ARC clinic notes.<br>• Infant medical records. | • The average age when a preliminary high-risk of CP or CP diagnosis was made, and age (/rate) of CP confirmation. |
| 6. To explore the experiences around communication and information sharing between families and health professionals. | • Semi structured interviews with families and with health professionals. | • Qualitative analysis of participant experience. |

#### Clinic/medical data

Quantitative data will be collected from medical records, PĒPI ARC clinic notes and through communication with local and regional health professionals. Data will include demographic and ethnicity information, indication for entry into the BPR pathway for CP, and completed early CP assessments including timing and results, diagnostic outcomes and referrals for monitoring and interventions.

Descriptive statistics will be used to report participant characteristics, recruitment rates, attendance rate in clinics as well as completion of diagnostic tools, referral for early intervention and surveillance. Analysis of Likert scale data from family, health professional and PĒPI ARC team questionnaires will also be analysed using descriptive statistics. Continuous data results will be reported using the median and interquartile range (lower and upper quartiles). Pearson’s Chi-squared or Fisher’s exact test will be used to compare the extent of implementation of the best practice guidelines between regions and ethnicities. A *p* value of <0.05 will be considered to be statistically significant.

#### Audit/records of communications (to health professionals and families)

Communication between the PĒPI ARC hub, families and health professionals will be captured via notifications to the PĒPI ARC email. These will be filed in the inbox according to themes throughout the duration of the study and then reviewed and analysed after study completion.

#### Questionnaires

Questionnaires will provide quantitative and qualitative information. Questions have been developed based on the theoretical framework of acceptability to evaluate the following constructs of acceptability: affective attitude, burden, ethicality, intervention coherence, opportunity costs, perceived effectiveness, and self- efficacy[16].

Families will be sent a link to the anonymous questionnaire within two weeks of attending the PĒPI ARC clinic, or at 5-6 months CGA if their infant is followed out in their local region. Display logic is embedded within the questionnaire to adjust the line of questions relating to in-person vs. virtual Hub attendance. All families will be asked to reflect on their experiences of communication, resources, and support.

Health professionals attending a PĒPI ARC clinic who are not a member of the regular PĒPI ARC team will be sent a survey within two weeks of the clinic appointment. Other health professionals not part of the PĒPI ARC team who have engaged with the Hub (through email audit trail review and being identified as being a primary person involved in the care of an infant) will be sent a survey invitation between 6-12 months of PĒPI ARC operating.

Members of the PĒPI ARC team will complete a specific team questionnaire focused on the acceptability of PĒPI ARC three months after clinic commencement. The data from the team-questionnaire will be analysed in real-time (i.e., within 4 weeks of capture) to allow for any changes to the study conduct if indicated. The questionnaire will be repeated at study endpoint to reassess for potential changes for service improvements.

Full analysis of all questionnaire data will occur at the end of the study period.

#### Semi-structured interviews

Semi-structured interviews will seek to capture the acceptability of PĒPI ARC[16] in addition to the experiences around communication of the diagnostic outcome, information sharing and resources and emotional supports provided. Questions will target the experiences of implementing the BPR within local and regional settings for health professionals, and support provided by the PĒPI ARC team. All semi-structured interviews will be offered to take place in a mode of the participant’s preference (face-to face at the site of the clinic Hub, or via videoconference). All interviews will be led by a researcher experienced in qualitative interviewing and will be audio recorded for transcription.

Families who have not opted out of being interviewed will be sent an email invitation once their infant is over the age of 6 months CGA. A staged review of recruitment of interviewees will occur after six months to ensure a cross section of infants has been captured i.e., regional and local, ethnic diversity especially representation of Māori and Pasifika, term and preterm infants, with/without likely diagnosis of CP.

Health professionals who have had interactions (i.e. received PEPI ARC clinic letters and GMA reports, email enquiries, education sessions etc.) with PĒPI ARC over the first 12 months, but are not part of the PĒPI ARC team, will invited to take part in an interview via email. Health professionals will be identified from attendance at in person clinics or zoom encounters, audit trail of emails and PĒPI ARC education/training sessions.

Interviews will be conducted until a saturation of experiences has been reached. The diversity of participants will be reviewed monthly thereafter, and targeted recruitment may be used. Qualitative content analysis will occur separately for family and health professional data but will occur concurrently to help determine any crossover of themes. Themes will be interpreted, discussed, and help inform the quantitative data. Data analysis will be completed using NVIVO software (QSR International 2021).

## ETHICS AND DISSEMINATION

This study has New Zealand Health and Disability Ethics Committees approval 2022 FULL 13434 and is registered on the Australian New Zealand Clinical Trials Registry (ACTRN12623000600640). Any modifications to the study protocol will be submitted as an amendment to the Ethics Committee and an update will be provide to the ACTRN registration. Informed written consent will be obtained from participants, and data will be de-identified for reporting. Health professionals completing anonymous questionnaires will indicate their consent by questionnaire completion, and those completing interviews will provide written consent. Results of the study will be published and disseminated (co-authored by investigators meeting ICMJE authorship criteria) in conference abstracts and presentations, peer-reviewed articles in scientific journals, the PĒPI ARC community and organisation newsletters and media releases. A de-identified dataset may be made available upon reasonable request but may be restricted by ethical limitations.

## DISCUSSION

This paper details the protocol for a feasibility study to ascertain if a multidisciplinary regional Hub, PĒPI ARC, can support equitable implementation of the NZ Early Detection of CP BPR for high-risk infants. Feasibility will be assessed for Acceptability and Demand among families and health professionals, and for Efficacy in relation to reducing the age of CP diagnosis and improving experiences around communication and information sharing between families and health professionals.

The study is limited to only infants with detectable risk-factors for CP admitted to Wellington NICU and who either live in Wellington or in one of its referral regions. As there is no control group, our ‘usual care’ infants with the same risk factors who decline to participate will not be followed and therefore cannot be compared to PĒPI ARC participants. Economic analysis of the effectiveness of PĒPI ARC for family and the health system is not being conducted.

The study has a number of strengths. The delivery of the Hub using two parts: a MDT face-to-face clinic and a regional virtual approach provides an opportunity to decease inequitable access to care for local and regional infants and families. Some regional centres may not have the experienced staff to deliver the BPR and the Hub will support and educate clinicians to ensure regional centres can, over time, deliver the recommended assessments and follow-up. The inclusion of qualitative feedback from both families and health professionals will provide depth of context to our findings.

## Data Availability

All data produced in the present study will be available upon reasonable request to the authors

## STUDY STATUS

Recruitment for the PĒPI ARC Hub opened to eligible participants in March 2023, with the first participant seen in the face-to-face clinic in June 2023. Participants will continue to be recruited until the end of February 2024 with the trial end date planned for September 2024.

## AUTHORS’ CONTRIBUTIONS

AAF and SW conceived the study design and AAF, SW, GK and MS the choice of outcome measures. AAF and SW acquired funding through the Cerebral Palsy Alliance. AAF, SU, AS, KB, SS, HF, and CB are PĒPI ARC team members managing all aspects of study implementation from NICU. AAF, SK, HF, SS and CB are members of the PĒPI ARC MDT clinic team. AAF, SW, AS, MS and SK conceptualised and led measurement of early detection; resources were developed by AS, SK, SU; GK and MS developed the questionnaires and interview framework; LW, GK and AAF developed and led the statistical plan for the study; SW provided expert contribution in feasibility study design and implementation. AAF, SW and GK completed the initial draft of the protocol manuscript. All authors have read, edited and approved the final manuscript.

## ACKNOWLEDGEMENTS

The authors would like to thank our Steering Committee, Advisory Committee and Whānau Hui who all contributed through co-design to the protocol. Specific support for ensuring Māori consideration was sought from Dr Paula King and Wyllis Korent and the development of the newborn passport into Te Reo Maori was made possible by Wyllis Korent. Pasifika engagement was supported by Janet Moananu.

## FUNDING STATEMENT

The work was funded by Cerebral Palsy Alliance (PRG01021), New Zealand Country Women’s Institute, and Health Research Council of New Zealand Research Activation Grant (22/696/A).

## COMPETING INTERESTS STATEMENT

The authors have no conflicts of interest to declare.

## COPY RIGHT/ LISENCE FOR PUBLICATION

The Corresponding Author has the right to grant on behalf of all authors and does grant on behalf of all authors, a non-exclusive licence on a worldwide basis to the BMJ Publishing Group Ltd to permit this article (if accepted) to be published in BMJ editions and any other BMJPGL products and sublicences such use and exploit all subsidiary rights, as set out in our licence

## Notes

### Competing Interest Statement

The authors have declared no competing interest.

### Funding Statement

The work was funded by Cerebral Palsy Alliance (PRG01021), New Zealand Federation of Women's Institute and Health Research Council of New Zealand Research Activation Grant (22/696/A).

### Author Declarations

Wellington Regional Hospital. The New Zealand Health and Disability Ethics Committees approved this study (HDEC:2022 FULL 13434).

